# Focal amplification of *FAL1*, an oncogenic enhancer lncRNA mapping to chromosome 1q is associated with dysregulated BMI1/p21 axis and an adverse event in intracranial ependymomas

**DOI:** 10.1101/2021.03.24.21254063

**Authors:** Prit Benny Malgulwar, Pankaj Pathak, Vikas Sharma, Pankaj Jha, Aruna Nambirajan, Madhu Rajeshwari, Manmohan Singh, Vaishali Suri, Chitra Sarkar, Mehar Chand Sharma

**Author notes:** **Address for Correspondence**: Mehar C. Sharma, M.D., F.R.C.Path, Professor, Department of Pathology, All India Institute of Medical Sciences, Ansari Nagar, New Delhi – 110029, India. Department of Translational Molecular Pathology, University of Texas-M.D. Anderson Cancer Center, Houston, Texas. National Institute of Neurological Disorders and Stroke, National Institute of Health (NIH), USA.

## Abstract

Gain of chromosome 1q locus is a common cytogenetic feature associated with intracranial ependymomas; however, candidate non-coding RNAs on this locus have not been identified. Recent studies have reported somatic copy number alterations for long non coding RNA (lncRNA) *FAL1/FALEC* residing on chromosome 1q to stabilize BMI1, an epigenetic repressor and PRC1 component, leading with to downregulation of p21/CDKN1A tumor suppressor gene. We aimed to study the role of *FAL1* in ependymomas, its association with 1q gain, BMI1/p21 regulatory axis and clinicopathological parameters. Using SNP array analysis (GSE32101), 31% (discovery cohort) and 63.8% (in-house cohort) showed amplification/gain of *FAL1* locus with higher prevalence in intracranial tumors. Copy number gain of *FAL1* locus was significantly associated with increased *FAL1* (p=0.003) and *BMI1* (p=0.007) levels, and reduced p21 (p=0.001) expressions. Interestingly, gain of *FAL1* locus and *FAL1* transcripts did not show any association with 1q gain or *RELA* fusions. Subcellular localization reported *FAL1* to be expressed in nuclear compartment in ependymomas. Chromatin immunoprecipitation-qPCR demonstrated *in-vivo* binding of BMI1 at p21 promoter locus with BMI1 target genes to be enriched in cell architecture related pathways. A 3-tier survival analysis between *FAL1* gain and increased expression levels of *FAL1* and *BMI1* correlated with poor outcome in our cohort. Ours is the first study demonstrating gain of *FAL1* locus and its interplay with the *BMI1*/p21 axis in intracranial ependymomas. Further studies into this epigenetic regulatory mechanism will unravel novel driver mutations in intracranial ependymomas

**Highlights:** Somatic variations in enhancer long non-coding RNA has been recently attributed for various clinical malignancies including cancers. Gain of 1q locus is a common cytogenetic variation observed in intracranial ependymomas. Our study has demonstrated, focal amplification of enhancer lncRNA mapping to Chromosome 1q, *FAL1/FALEC*, to be involved in oncogenicity/ progression of ependymomas. Moreover, our data suggests a positive association with BMI1 (a PRC1 component) with *FAL1* levels, indicating downregulation of BMI1 target gene involved in cell cycle, p21. Furthermore, a 3-tier prognosis analysis (between *FAL1* gain, *FAL1* and *BMI1* expressions) suggests a negative survival outcome. Our study highlights the importance of somatic variation in non-coding genome with ependymoma survival.

## Introduction

Ependymomas are neuroepithelial tumors that recapitulate the ependymal cells lining the ventricles and spinal canal^1^. They are the third most common central nervous system tumor in children^2^. Despite advances in diagnostics and therapeutics, there has been little improvement in the outcome of these tumors. Age, site and extent of resection are the most consistently identified factors determining outcome, while WHO histological grade by itself does not always correlate with survival^3,4^. There are few established prognostic markers, of which gain of chromosome 1q has been consistently identified as a poor prognostic marker in many studies^3,5,6^. 1q gain is a common event in ependymomas, identified in 13-33% of all ependymomas^5,7,8^ and more frequent in children, infratentorial location and tumors with anaplastic histology^5,8-11^. A recent molecular subclassification identified nine molecular subgroups of which 1q gain was observed to be frequent event (25%) in the aggressive posterior fossa and supratentorial molecular subgroups^12^. In addition to paediatric ependymomas, gain of 1q has also been demonstrated as an adverse prognostic factor in neuroblastoma, Wilms tumor and Ewing sarcoma perhaps indicating a wider role for the involvement of 1q in the progression of paediatric tumours^13-16^.

Long non-coding RNAs (lncRNAs) are anti-sense RNA derived from protein-coding genes, ultra-conserved regions and introns ^17,18^. They range from 200 nucleotides to ∼100 kb and functions as enhancer-like elements to various proteins^19^. A wide range of lncRNAs have been reported in literature, of which *ANRIL, HOTAIR, HOTTIP, MEG3* have demonstrated oncogenic potential and serve as prognostic biomarkers in gliomas, medulloblastomas and spinal ependymomas^20-22^. Genome-wide study on somatic copy number alterations for lncRNAs (long non coding RNAs) has identified a novel oncogenic enhancer lncRNA, *FAL1, (*<u>a</u> focally <u>a</u>mplified lncRNA on chromosome <u>1</u> (ENSG00000228126)) or *FALEC*, residing on chromosome 1q21.2, to be associated with poor outcome in ovarian cancer^23^. Series of functional experiments demonstrated that *FAL1* interacts with and modulates the activity of epigenetic repressor *BMI1* with resultant downregulation of multiple genes including tumor suppressor gene *CDKN1A*/p21^23^. Recent literature further suggests dysregulation of FAL1 in multiple cancers (including non-small lung cancer, esophageal cancer, thyroid cancer, hepatocellular cancer, prostate cancer, cervical cancer, and oral squamous cell carcinoma) and other diseases including (Hirschsprung’s disease and diabetic arteriosclerosis), indicating *FAL1* as potential diagnostic and prognostic marker^24^.

The genomic region specific for 1q gain in ependymomas includes 1q21–q31, 1q21.3 –q23.1, 1q22–q31, 1q31.1–q31.3, 1q31– q32 and 1q41– 1qter^6,7,25^, of which high-level gain has been found to be restricted to 1q21 to q31^26^. The role of these amplicons remains unclear, and although a few potential candidate coding genes have been identified in this region of gain^7,27,28^, none of them directly relate to ependymoma tumorigenesis, relapse or patient outcome. In the light of evidence for the oncogenic mechanism of 1q gain involving *FAL1* enhancer lncRNA, we aimed to study the role of *FAL1* in pediatric ependymomas, its association with 1q gain, BMI1/p21 regulatory axis and clinicopathological parameters.

## Materials and Methods

### SNP array analysis

SNP array analysis was performed using publically available data on GEO for pediatric ependymoma using GSE32101^5^. GSE32101 contains data for 42 pediatric ependymoma which has been run on Affymetrix 500K SNP array platform.

### Sample collection

The study was a retrospective design and ethically approved by the Institute Ethics Committee (Ref No: IESC/T-211/05/05/2015). Fresh tumor samples from patients operated in the Department of Neurosurgery are collected from the operation theatre at the time of surgery and consent was taken from the patients/guardians. Portions of resected tumors are snap-frozen in liquid nitrogen and stored at -80 °C until use, and the remaining tissue is fixed in 10% buffered neutral formalin and paraffin-embedded for routine histopathology and immunohistochemistry. Cases diagnosed as ependymomas between 2003 and 2016 were retrieved from the archives and corresponding Hematoxylin and Eosin (H&E) stained slides were reviewed for reconfirmation of diagnosis by three neuropathologists (MCS, MR & AN) according to the 2016 update of the World Health Organization (WHO) classification of CNS tumors.

### Realtime qPCR studies for gain of *FAL1* locus

Eight to ten 10µm-thick serial sections were collected from each formalin-fixed paraffin-embedded tumor tissue block. Deparaffinization was followed by Proteinase K digestion. For DNA isolation Recover All Total Nucleic Acid Isolation kit (Ambion) was used as per manufacturer’s instructions.

Gain of *FAL1* was sought for by using primers spanning *FAL1* locus and normalized using primers for genomic *GAPDH*. Additionally, DNA extracted from FFPE tissue sections of normal brain (ependymal scrapings from lateral ventricles and fourth ventricle) were used to represent control samples. Realtime-qPCR was performed using Syber-green using LC480 Real-Time PCR System (Roche Diagnostics, Switzerland) and copies detected more than 4 fold change/copy was considered as positive. The primer sequences used for copy number analysis are provided in **Supplementary Table**-1.

### Gene expression analysis for FAL1, BMI1 and CDKN1A/p21

Total RNA was isolated using mirVana™ miRNA Isolation Kit (M/S Ambion, Life Technologies, USA) as per manufactures protocol. One µg of total RNA was reverse transcribed using Superscript VILO cDNA Synthesis Kit (M/S Invitrogen, Life Technologies, USA). Quantitative Realtime PCR (qPCR) was performed using Syber-green with LC480 Real-Time PCR System (Roche Diagnostics, Switzerland). The differences in expression between patients and controls (delta Ct) were calculated using the comparative method and the level of *FAL1, BMI1* and *CDKN1A*/p21 fold change was calculated using 2−ΔΔCt method. The primer sequences for the transcripts analyzed are provided in **Supplementary Table**-1.

Additionally, 4 gene expression microarray dataset of ependymomas: GSE21687^29^, GSE27279^11^, GSE50385^30^ and GSE64415^12^ and normal brains (GSE66354) were downloaded from GEO repository and analyzed in-house as well as using R2 software (https://hgserver1.amc.nl/cgi-bin/r2/main.cgi)

### *RELA* fusion analysis

Detection of *RELA* fusion transcripts were performed using cDNA as reported by Malgulwar PB, et al^31^ and bidirectional sequencing was performed using the BigDye Terminator v3.1 Cycle Sequencing Kit (Applied Biosystems, Courtaboeuf, France) using the ABI 3130xL sequencer (Applied Biosystems, Foster City, CA, USA).

### 1q gain analysis

Gain of 1q locus was performed as per Rajeshwari M, et al^8^ using dual colour interphase fluorescence in situ hybridization (FISH) probe for test locus 1q25 (spectrum green labelled) and for control locus 1p36 (spectrum orange labelled). Signals were scored in 100 non-overlapping, intact nuclei under oil immersion. Tumor signals were scored as gain when at least 10% of cells showed three or more signals of the test probe^3,7^, or if the ratio of test probe–control probe was more than one^6^.

### Subcellular fractionation

Subcellular fractionation for ependymoma tissues was carried out using the PARIS kit (M/s. Invitrogen) following manufacturer’s instructions. After isolating the nuclear and cytoplasmic RNAs from each subcellular compartment, RNA was reverse transcribed and quantified as described above. Using qRT-PCR, expression levels for FAL1 were measured independently in the cytoplasmic and nuclear fractions and relative expression was plotted using U6 snRNA and GAPDH as controls.

### Immunohistochemistry for MIB1

Immunohistochemical studies were performed on 5-µ-thick formalin-fixed, paraffin-embedded tumour sections using antibodies directed against MIB-1(Dako, Denmark; 1:200). Labelled streptavidin biotin kit(Universal) was used as a detection system(Dako, Denmark). MIB-1 labelling index was calculated as percentage after counting 1000 tumor cell nuclei in different hot spots.

### Chromatin Immunoprecipitation-qPCR

Chromatin Immunoprecipitation-qPCR were performed as per Sharma V, et al^32^ with some minor modifications. Briefly, frozen ependymoma tissues were thawed on ice and chopped into small pieces and was cross-linked in 1% formaldehyde in PBS for 10 min. The crosslinking reaction was stopped using 100 ul of 125 mM glycine. The chromatin was next fragmented using Diagenode Bioruptor plus at high power settings (as per manufacturers guidelines). Fragmented chromatin was immunoprecipitated using anti-mouse BMI1 (M/s Abcam) and anti-mouse IgG (M/s Abcam) antibody as per Low cell ChIP kit guidelines. After overnight incubation, immunoprecipitated and input DNAs were purified in 40 µl of water with IPure kit following the manufacturer’s instructions (M/s Diagenode). qPCR reactions were performed on 3 µl of DNA in a LC480 system (Roche) using Syber green dye for p21 promoter sites and ACTB. The sequences of the primers used for detection is provided in **Supplementary Table-1**. Enrichment was expressed as the percent input by using the following formula: Percentage of total Input=100×2^[Ct (ChIP) – (Ct input – log2 (input dilution factor)].

### Chromatin Immunoprecipitation-seq (ChIP-seq) data for BMI1

Genomic targets for BMI1 was obtained from Gargiulo G, et al.^33^ which was performed on 5 GBM tissue samples.

### Statistical analysis

Kaplan–Meier survival analysis was used to estimate the survival in the present cohort. Cox proportional hazard, Pearson correlation, ANOVA and T-test were performed using SPSS version 11.5 for Windows. GraphPad Prism version 5.0 for Windows and Microsoft Excel was used for constructing scatter and box plots. Additionally, clinical relevance of BMI1 expression was evaluated in an independent ependymoma cohort (GSE27287) [11]. In all test, 2-sided p-value of less than 0.05 was considered as significant.

## Results

### Clinicopathological features of selected ependymoma samples

A total of 47 ependymomas were included in our study **(Supplementary Table-2)**. *C11orf95-RELA* fusions were detected in 71.4% (15/21) of supratentorial ependymomas, identifying this sub-set as the ST-EPN-*RELA* molecular subgroup, with a median age of 9 years (range: 1 to 29 years), male preponderance (2:1 male:female ratio) and predominantly Grade III histology. The remaining six *RELA*-fusion negative ependymomas were designated as the ‘ST-*RELA(*-) subset’. These tumors, showing a wider distribution of age (median 16.5 years, range 2-59 years), likely belong to the ST-EPN-*YAP1*. Due to non-availability of DNA methylation assays, we attempted to categorize posterior fossa ependymomas into the molecular subsets, viz. PF-EPN-A and PF-EPN-B based on age. 80% (12/15) of the PF ependymomas included occurred in children (median age at diagnosis 10.5, range 1.5 to 16 years), roughly representative of the PF-EPN-A subgroup, while the remaining occurred in adults (median age 33, range 33-46), representing the PF-EPN-B subgroup. The spinal ependymomas were all of Grade II/III histology, representative of the SP-EPN subgroup, occurring at a median age of 22.5 years (range 9 to 55 years). FISH for 1q locus gain was performed in 20 cases and gain was observed in 4/9 supratentorial ependymomas, including 2/4 *RELA* fusion positive ependymomas, 4/9 pediatric PF ependymomas, and 1/2 adult PF ependymomas.

### Analysis of FAL1 locus gain in ependymomas

*FAL1*, a focally amplified lncRNA on chromosome 1 (FAL1, ENSG00000228126), is a long non-coding RNA gene located on chromosome 1q21.2^34^. Because gain of 1q locus is frequently observed cytogenetic hallmark of intracranial ependymomas, we hypothesized that *FAL1* may represent a potential gene of interest in this locus. As an initial analysis (discovery cohort), we performed a bioinformatics analysis for SNP array data for 42 ependymomas from GEO (GSE32101) performed on Affymetrix 500K SNP arrays, specifically for 1q locus. Interestingly, 31% (13/42) of cases showed gain of *FAL1* locus **(Figure-1a)**.

**Figure 1:**
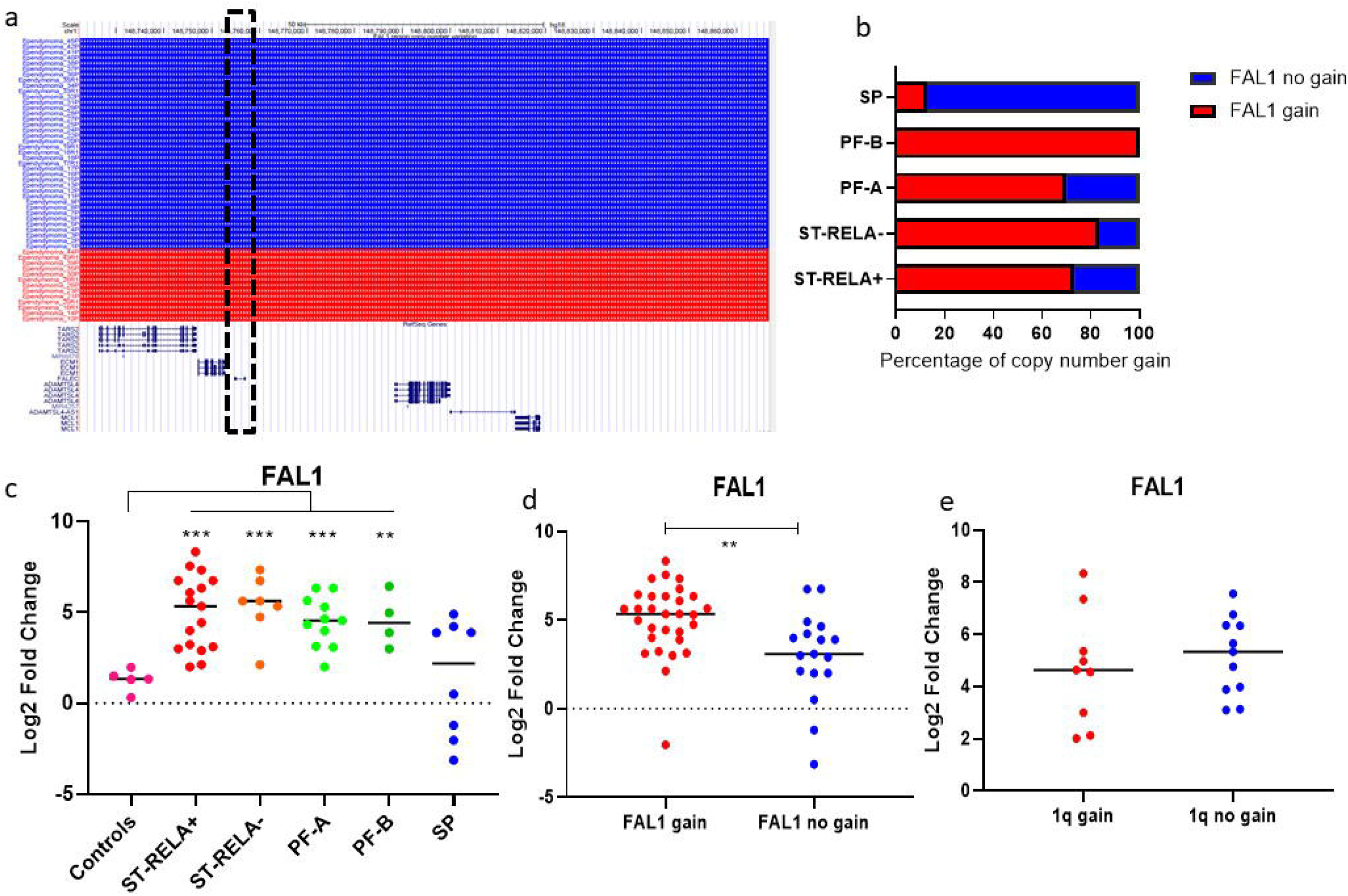
*FAL1* copy number gain and expression analysis in ependymomas: UCSC visualization screenshot for SNP array data-GSE32101, highlighting 1q21.2 locus for *FAL1* in black box. Red color indicates gain of the genomic segment, while blue color indicates presence of normal copies (a). (b) Validation of *FAL1* gain in different molecular subgroups of ependymomas in in-house cohort. qPCR for *FAL1* expression levels in (c) different molecular subgroups, (d) *FAL1* gain and (d) 1q status. Significance representation-<0.0001-****, <0.001-***, <0.001-**, <0.01-*.

To validate these findings, we performed qPCR for copy number gain of *FAL1* locus in our cohort (validation cohort). 63.8% (30/47) cases showed copy number gain of *FAL1* locus (≥4 copies), with higher preponderance in intracranial location i.e. 74% (29/39) as compared to spinal i.e. 12.5% (1/8) **(Figure-1b**). Within the intracranial subsets, there was no correlation observed with age, gender or histological grade. FAL1 gain was observed across the different intracranial ependymoma subgroups with ST-*RELA+* (73.3%, 11/15), ST-*RELA-*(83.3%, 5/6), PF-A (70%, 7/10) and PF-B (100%. 3/3). Interestingly, there was no correlation of *FAL1* gain with 1q gain, being observed in 56% and 73% of ependymomas with and without 1q gain respectively (p=0.6).

### Gain of *FAL1* locus is associated with overexpression of FAL1 expression

We performed Realtime-qPCR analysis to quantify *FAL1* gene expression in ependymomas. The mRNA levels of *FAL1* were significantly upregulated in intracranial molecular subgroups as compared to controls and strongly associated with FAL1 gain (p<0.001) **(Figure-1c and 1d)**. There was no correlation with 1q gain status **(Figure-1e)**. Notably, 50% of spinal ependymomas showed increased FAL1 expression without corresponding FAL1 gain, suggesting mechanisms other than genomic amplification may lead to overexpression in this subset as observed previously^23^.

### Gain of *FAL1* locus is associated with increased expression of BMI1, a Polycomb Repressive Complex (PRC) Molecule

Since the gain of *FAL1* locus correlates with *FAL1* expression in intracranial ependymomas, we further analyzed the expression of *BMI1* gene, a known collaborator of *FAL1* and a component of PRC1 complex. Gene expression analysis across 4 different ependymoma cohorts confirmed overexpression of *BMI1* genes as compared to controls (p<0.001) **(Supplementary Figure-1a)** with favorable expression in PF-A as compared to other molecular subgroups of ependymomas **(Supplementary Figure-1b)**. In-house validation of *BMI1* expression using Realtime-qPCR confirmed our in-silico analysis **(Figure-2a)**. On the basis of molecular subgroups, the expression of *BMI1* was higher in PF-A among all subgroups. There was no correlation with 1q gain **(Supplementary Figure-1c)**.

We next correlated the expression of *BMI1* with copy number gain of *FAL1* locus and *FAL1* expression. A significant association was observed between *BMI1* and copy number gain of *FAL1* locus (Pearson correltion:0.53) and between *BMI1* and *FAL1* expression (Pearson correlation:0.40) **(Figure-2b and 2e)**, indicating *FAL1* could be involved in stabilizing *BMI1*, as reported by Hu X, et al (2014).

**Figure 2:**
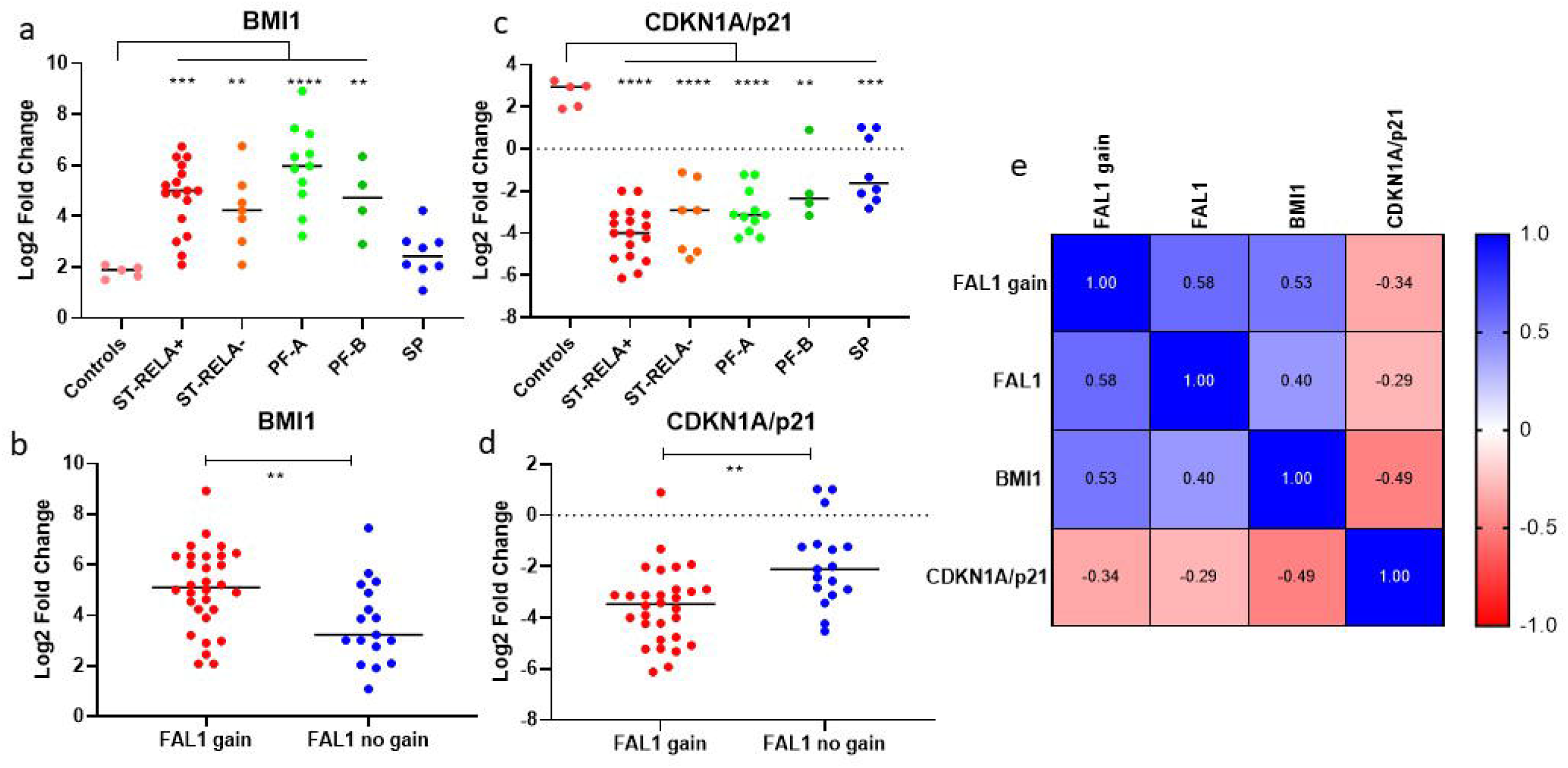
Expression for BMI1 and CDKN1A/p21 in ependymomas and their association with FAL1: BMI1 and CDKN1A/p21 expression levels in different molecular subgroups (a and c) and with *FAL1* status (b and d). Heatmap for correlation matrix for different variables (e). Significance representation-<0.0001-****, <0.001-***, <0.001-**, <0.01-*.

### Downregulation of *CDKN1A*/p21 is mechanistically linked to BMI1-*FAL1*

Among the various target genes common to both *FAL1* and BMI1, Hu X et al. demonstrated that the expression of tumor suppressor gene *CDKN1A/p21* is significantly downregulated by *FAL1. FAL1* is also involved by stabilizing BMI1 and thereby increasing H2AK119 ubiquitination at the promoter site of *CDKN1A*^23^. On analyzing the expression status of *FAL1* target genes by using 4 publicly available gene expression profiling data for ependymomas, we found that expression of *CDKN1A*/p21, a potent cyclin-dependent kinase inhibitor and an important component of p53/RB pathway, was significantly downregulated in ependymomas as compared to controls (p<0.001) **(Supplementary Figure-2a)** and among the posterior fossa molecular groups of Pajtler KW et al. ^12^ **(Supplementary Figure-2b)**. In our cohort, we confirmed the down regulation of *CDKN1A*/p21 expression as compared to controls (p<0.001) **(Figure-2c)**. On basis of molecular subgroups, the expression of CDKN*1A*/p21 was downregulated in ST-EPN-*RELA* in our cohort as well as in the Pajtler cohort^12^. In addition, we observed a significant negative correlation of *CDKN1A* with *FAL1* copy number gain (Pearson correlation: -0.34), *FAL1* expression (Pearson correlation: -0.29) and *BMI1* expression (Pearson correlation: -0.49) **(Figure-2d and 2e)**. A similar negative association between *BMI1* and *CDKN1A* levels was also observed in (Pearson correlation: - 0.41) in published datasets of Witt H, et al. (2011) **(Supplementary Figure-2c)**. There was no association between *CDKN1A* levels and 1q gain (**Supplementary Figure-2d)**.

Hu X et al demonstrated that BMI1 occupancy and H2AK119 ubiquitination of the promoter region of target genes, particularly *CDKN1A*, decreased following *FAL1* knockdown in ovarian cancer cells. We hypothesized that the decreased levels of *CDKN1A*/p21 in ependymomas may be due to a similar epigenetic silencing by BMI1-mediatied H2AK119 ubiquitination at *CDKN1A*/p21 promoters. To confirm this, we performed Chromatin immunoprecipitation-qPCR using BMI1 antibody on 3 ependymoma tumor tissue sample (2 PF-EPN-A and 1 ST-EPN-RELA). The ChIP-qPCR data analyses revealed enrichment of the *CDKN1A*/p21 promoter region with BMI1 suggesting BMI mediated epigenetic silencing of CDKN1A in ependymomas as well **(Figure-3a)**.

**Figure 3:**
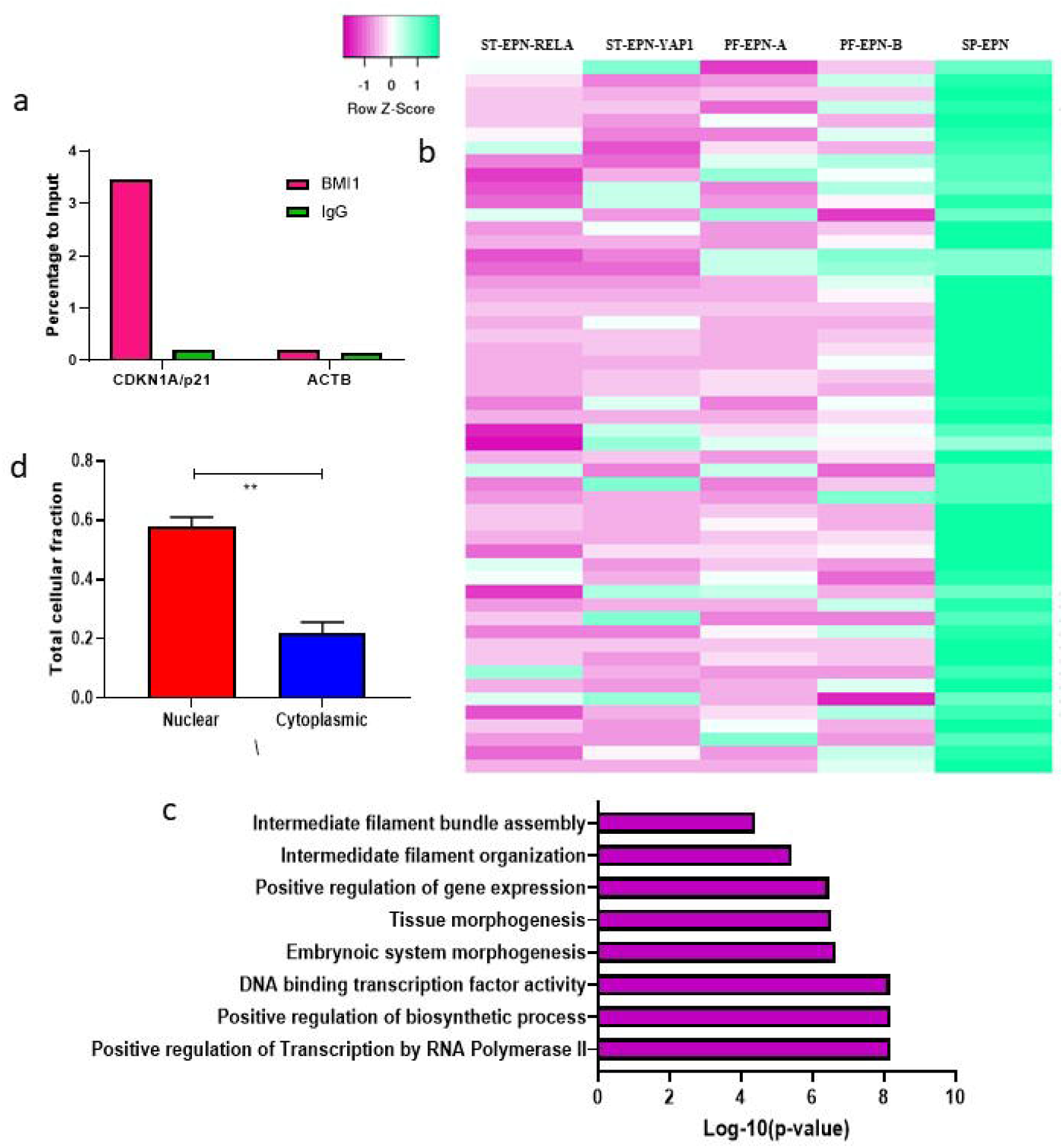
*FAL1/BMI1* target gene analysis and *FAL1* localization: ChIP-qPCR analysis for BMI1 occupancy at CDKN1A/p21 promoter in ependymoma tissues (a). Heatmap for BMI1 target genes in Grade II and III molecular subgroups of ependymomas (b). Gene ontology analysis for downregulated BMI1 target genes in intracranial tumors (c). RNA localization experiments performed using qPCR for *FAL1* (d). Significance representation-<0.0001-****, <0.001-***, <0.001-**, <0.01-*.

### *FAL1-*BMI1 association downregulates critical genes and pathways, promoting ependymomagenesis

As BMI1 is associated with downregulation of several tumor suppressor and cell cycle regulators via H2AK119 ubiquitination process, we reasoned that BMI1-FAL1 corroboration may affect genome-wide modifications in transcription. To test this hypothesis, we examined the BMI1-dependent genes in ependymomas using ChIP-seq profiling data in Gargiulo G, et al. ^34^ and annotated their expression status in various molecular subgroups, both intracranial and spinal, as in Pajtler KW, et al. ^12^ **(Figure-3b)**. Genes involved in cell architecture (*DAAM1, DSP, NEFL, TMOD1*) and HOX clusters including *HOXA13, HOXB2, HOXB5, HOXB7* and *HOXC8* were found to be downregulated. Upon Gene Ontology annotation, pathways related to transcription (p=6.42⨯10^−9^), biosynthetic (p=6.57⨯10^−9^) process were significant. Surprisingly, Intermediate filament assembly (p=3.98⨯10^−5^) and organization (p=3.86⨯10^−6^) were found enriched **(Figure-3c)**.

### *FAL1* transcripts are localized in nuclear compartment

In order to elucidate the potential oncogenic mechanism of *FAL1* transcripts, we investigated the subcellular localization of *FAL1* by qRT-PCR in 3 ependymoma tissue samples. We found that *FAL1* lncRNA is primarily located in the nuclear compartment of ependymoma tumor cells **(Figure-3d)** supporting previous observations that *FAL1* exerts its regulatory function at the nuclear level^23^.

### FAL1 gain is associated with high proliferation rate

CDKN1A/p21 is a potent cyclin-dependent kinase inhibitor and an important component of p53/RB pathway, and our results suggest that the oncogenic potential of *FAL1* is likely related to the downregulation of this tumor suppressor. We assessed the proliferation rate in ependymomas using MIB1, and found that high MIB1 L1 (≥10%) showed a significant association with gain of FAL1 locus (p=0.035) **(Figure-4a)**.

**Figure 4:**
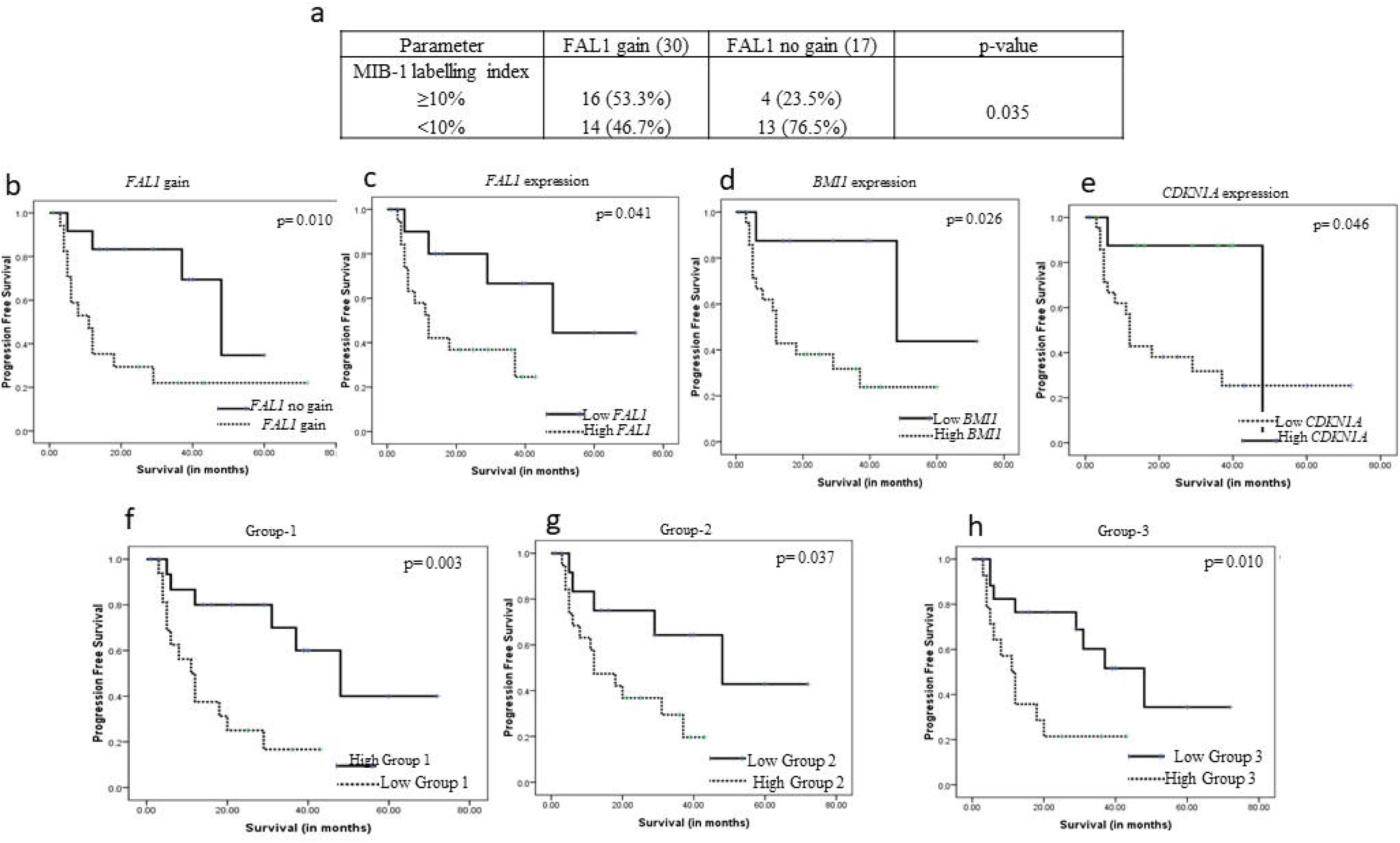
Correlation of *FAL1* with proliferative and survival status: *FAL1* copy number status with MIB1 proliferative indices (a). Univariate Progression Free survival plots for *FAL1* gain (b), *FAL1* (c), *BMI1* (d), *CDKN1A* (e) expression. 3-tier expression survival analysis for *Group-1*-Cases with *FAL1* gain and high *BMI1* expression (f); *Group-2*-Cases with high *FAL1* and high *BMI1* expression (g); *Group-3* Cases with *FAL1* gain and high *FAL1* and *BMI1* expressions (h).

### Survival analysis

*FAL1* gain (p=0.01), high *FAL1* transcripts (p=0.04), high *BMI1* transcripts (p=0.01) and low *CDKN1A*/p21 transcripts (p=0.01) **(Figure-4b-4e)** associated with shorter progression free survival (PFS) on univariate analysis. We confirmed our survival analysis in an independent external ependymoma cohort (GSE27287) and found high *BMI1* (p=0.040) and low *CDKN1A*/p21 (p=0.017) expression showing a trend to shorter PFS **(Supplementary Figure-3)**.

Next, we determined the association of combinations of three biomarkers: *FAL1* gain, *FAL1* expression, and *BMI1* expression with patient outcome as follows: *Group-1*-Cases with *FAL1* gain and high *BMI1* expression; *Group-2*-Cases with high *FAL1* and high *BMI1* expression; *Group-3* Cases with *FAL1* gain and high *FAL1* and *BMI1* expression **(Figure-4f-4h)**. Interestingly, all the three groups showed a significant trend towards poor survival, indicating that this 3-tier combination of genomic gain (*FAL1*), post-translation modification (*FAL1* expression) and epigenetic dysregulation (*BMI1* expression) has a deleterious effect on survival of patients.

## Discussion

Gain of chromosome 1q is a common cytogenetic alteration observed in various tumors including sarcomas^35^, neuroblastoma^16^, Wilms tumor^36^ and atypical meningioma^37^. In ependymomas, gain of 1q locus has been reported to be a marker of recurrence-free and overall survival in numerous studies^3,5,7,12^, although few studies do contradict these findings^6,8^. In a recent multi-institutional study of 500 ependymomas, 1q gain emerged as an independent predictor of poor overall and progression free survival^12^. The oncogenes located at 1q locus are yet to be identified in ependymomas and although a few candidate genes have been studied, the oncogenic potential of the non-coding genes located at 1q locus have not been explored previously. In the present study, we aimed to evaluate the role of *FAL1*, an enhancer lncRNA, located at 1q21.2 locus as a candidate oncogene in ependymomas. For the first time, we report the frequent occurrence of *FAL1* locus amplification in intracranial ependymomas. Although the copy number gain of *FAL1* locus occurred far more frequently in intracranial ependymomas than 1q gain, and showed poor correlation with the latter, we further unraveled the functions of *FAL1*. As previously described, our study confirmed its association with epigenetic regulator *BMI1* and their role in the downregulation of the tumor suppressor gene *CDKN1A/p21*, thus unveiling *FAL1* as a potential oncogene and therapeutic target in intracranial ependymomas^23,24^.

LncRNAs are now emerging as crucial participants in cancer biology. Several genome-wide profiling studies for lncRNAs have revealed differential expression patterns in normal tissues and cancers^38^. Recent literature suggests an important role for lncRNAs in the reprogramming of the chromatin architecture of cancer cells, thereby modulating the cancer epigenome and serving as potential targets for cancer diagnosis and therapy^17,18^. Genes coding for lncRNAs can be found in intergenic or intronic regions, or at overlapping or antisense loci adjacent to protein-coding genes, where they are postulated to exert regulatory functions. Somatic copy number alterations (SCNAs) leading to altered expression of lncRNAs has been previously reported in neuroblastoma where gain of 17q locus is linked to expression of *ncRAN* (non-coding RNA expressed in aggressive neuroblastoma), located at 17q25.1 locus^39^. In another recent study on colonic carcinomas, Ling H et al^40^ reported activation of a lncRNA, *CCAT2*, resulting from SNPs within the 8q24 locus, that promotes tumor growth and metastasis by upregulation of WNT signaling.

LncRNAs are abundantly expressed in the brain compared to other regions^41^. However, the vast majority of lncRNAs in the brain have not yet been functionally characterized. Some of the known oncogenic lncRNAs reported in gliomas are H19, which is highly expressed in gliomas and has potential to bind with c-myc to drive tumorigenesis^42^. Li R, et al. ^43^ has classified glioma based on lncRNA signature subgroups which was found to be strongly associated with disease outcome. Other oncogenic lncRNAs include MALAT1^44^ and HOTAIR^45^ whose increased expression has been correlated with poorer overall survival.

*FAL1* is a focally amplified lncRNA, located on 1q arm of chromosome 1 and has 2 coding exons^34^. *FAL1* locus gain is a common event in epithelial cancers, occurring in nearly 50% of carcinomas, while is rare in CNS tumors such as medulloblastomas and astrocytomas, occurring in less than 20% of them^23^. In the present study, we demonstrated gain of FAL1 locus in approximately 63.8% of ependymomas, with incidence increasing to 72% among those arising in intracranial locations. The *FAL1* locus gain corresponded with a simultaneous increase in *FAL1* lncRNA transcripts within the nuclei of ependymoma tumor cells, indicating that the genomic amplification is functional and is not merely a passenger event^23^. Another observation in our study was the frequent overexpression of *FAL1* RNA in the spinal ependymomas, in which FAL1 copy number gain was not seen. Hu X, et al^23^ et al also noted that several cell lines without genomic amplification of *FAL1* did overexpress *FAL1* mRNA, suggesting that FAL1 overexpression is common in malignancies and may be caused by mechanisms other than genomic amplification.

*FAL1* gene is located at 1q21.2 and in our study, we observed that there was surprisingly poor correlation between gain of 1q25 locus (via FISH assay) and *FAL1* locus gain or *FAL1* RNA transcript levels (via qRT-PCR). Among cancer causing genomes, somatic copy number alterations involving small foci of the chromosome are more likely to harbor cancer causing genes rather than those involving a large fragment or an arm of the chromosome^46^. In particular Hu X et al^23^ noted that *FAL1* gene often resides at a significant focal amplicon (Q<0.25) on chromosome 1q21.2 in epithelial cancers. Thus, it is likely that the focal somatic copy number alterations leading to *FAL1* gain may be too localized to be detected by FISH. Thus, it would be advisable to assess the genomic copy number gain via qPCR rather than FISH assay for assessing *FAL1* gain. FAL1 gain and increased FAL1 expression associated with a poorer progression free survival in our cohort and similar poor prognosis has been previously observed in ovarian carcinomas and other cancers^23,24^.

*BMI1* is a member of Polycomb Repressive Complex 1 (PRC1), serving to stabilize the whole PRC1 complex that represses the transcription of various genes by histone modifications^47^. Particularly, Hu X et al demonstrated that BMI1 protein increases the ubiquitination levels of H2AK119 on the promoter regions of its target genes. BMI1 is an important regulator of self-renewal in neural stem cells^48^ and is critical in establishing stem-cell like traits in embryonic stem cells (ESCs) ^47^. In cancers, BMI1 acts as an oncogene by upregulating c-myc^49^ and down regulating tumor suppressors: *p16INK4a* and *p19ARF*^50^. Few studies have reported increased expression of *BMI1* in gliomas^51^ and medulloblastomas^52^. Farivar S et al^53^ reported 8.64 fold increase in the expression of *BMI1* in ependymomas as compared to other pediatric brain tumors (medulloblastoma, astrocytomas, Primitive Neuro Ectodermal Tumors (PNET), gangliogliomas, and oligodendrogliomas). High expression levels of BMI*1* in ependymomas was also reported by Baxter PA, et al^51^. BMI1 is a marker of neural stem cells and is involved in maintenance of neural stem cell self-renewal and neural development^48^. Aizawa T, et al^54^ reported frequent expression of BMI1 protein in an ependymoma cell line, that was developed using mouse neural stem cells transfected with human BK polyomavirus. In our study, gene expression for *BMI1* was elevated more in intracranial ependymomas as compared to their spinal counterparts. Studies on ependymomas have previously shown that cancer stem cell-like phenomenon is enriched in intracranial locations as compared to spinal^31^, possibly explaining the elevated expression levels of BMI1 in intracranial locations. The exact molecular mechanism behind *BMI1* upregulation remains unclear. Similar to our findings, Hu X et al^23^ also found a close concordance between the expression levels of *FAL1* and *BMI1*, and demonstrated that *BMI1* regulate target genes related to cell architecture and HOX clusters. However, the mRNA levels of *BMI1* were not dependent on *FAL1* expression. Rather, they demonstrated that *FAL1* lncRNA interacts with and stabilizes BMI1 protein, increasing its half-life and considerably augmenting its oncogenic potential. Functional knock down assays need to be performed to ascertain whether BMI1 expression is dependent on FAL1 expression and the possibility of common upstream regulators.

*FAL1*/BMI1 interaction has been shown to represses various genes involved in cell proliferation, cellular movement and protein sorting, particularly *CDKN1A/p21*^23^. We also demonstrated downregulation of *BMI1* targets genes in intracranial ependymomas across all molecular subsets. Among the various genes that were repressed, *CDKN1A/p21* plays a central role in arresting cell cycle and was selected as a candidate tumor suppressor gene^55^. It behaves as a universal inhibitor of cyclin-dependent kinases (CDKs) and plays critical roles in G1/S and G2/M cell cycle transitions^55^. By chromatin pull down study, we demonstrated that BMI1 protein binds to the promoter region of *CDKN1A/p21* in ependymoma tumor cells, in support of previous observations that BMI1 protein binds to the promoter region of *CDKN1A/p21*, increases the ubiquitination of H2AK119 and represses p21 expression^23^. Binding of BMI1 to *CDKN1A/p21* promoter has been reported in human glioblastomas^48^. P21 repression leads to activation of cell cycle, as reported by Jacobs JJ et al^50^ and Molofsky AV, et al. ^48^. A recent study by Tzaridis T, et al^56^ demonstrated that administration of low dose of Actinomycin-D in PF-EPN-A and ST-EPN-RELA cell lines, resulted in an increase in the expression of p21 protein. Actinomycin appears to be feasible treatment option that needs to be explored further in intracranial ependymomas with FAL1/BMI1/p21 dysregulation.

The dissection of the prognostic relevance for *FAL1* gain and its associated molecules furthers strengthens our data from clinical perspective. We report a unique 3-tier combination of genomic gain (*FAL1*), a non-coding RNA (*FAL1*) and a epigenetic regulator (*BMI1*) to be a poor prognostic factor in ependymomas. In the present study, for the first time we highlighted the possibility of prognostically stratifying ependymomas by evaluating the status of these molecules by using a method (qPCR) that is easily applicable in daily practice. However, the number of cases with follow-up data in the present study is limited, hence the combinational prognostic significance of these three molecules must be validated in a larger cohort of ependymomas.

Our study is the first to demonstrate the frequent gain of focally amplified lncRNA (FAL1) in ependymomas and its oncogenic role in repressing p21 by an association with epigenetic regulator (BMI1). The study needs to be extended to *in-vitro* and *in-vivo* mouse models of ependymomas to validate our findings and evaluate therapeutic prospects of FAL1 targeting.

## Conclusion

Genomic amplification of *FAL1* is a frequent event in intracranial ependymomas, and is significantly correlated with *FAL1* and *BMI1* expressions. There exists a mechanistic relationship between BMI1 and *CDKN1A*/p21. There was no correlation with molecular subsets within intracranial locations or 1q gain. A 3-tier system associated with poor prognosis in ependymomas. Further evaluation of *FAL1* locus and its associated molecular features may serve to identify potential therapeutic targets and prognostic markers.

## Data Availability

SNP array analysis was performed using publically available data on GEO for pediatric ependymoma using GSE32101. The list of 47 ependymomas with their genetic alterations can be found in Supplementary Table-2

## Contribution of listed authors

**Conception and design:** Prit Benny Malgulwar, Mehar Chand Sharma. **Performed the SNP array, copy number gain analysis:** Prit Benny Malgulwar, Pankaj Pathak, Pankaj Jha. **Gene expression experiments and data analysis:** Prit Benny Malgulwar. **ChIP-qPCR and RNA localization experiment**: Prit Benny Malgulwar, Vikas Sharma. ***RELA* fusion and FISH analysis for 1q gain:** Prit Benny Malgulwar, Madhu Rajeshwari, Aruna Nambirajan. **Contributed reagents/ materials/analysis tools:** Mehar Chand Sharma, Chitra Sarkar, Vaishali Suri, Manmohan Singh. **Wrote the paper:** Prit Benny Malgulwar, Mehar Chand Sharma.

## Acknowledgment and Funding

The authors are thankful to Neuro Sciences Centre, AIIMS and Science and Engineering Research Board (SERB) (EMR/2016/003365) for financial support; all consultants from the Departments of Pathology and Neurosurgery, AIIMS; all technical staff from the Neuropathology laboratory, AIIMS and Indian Council of Medical Research(ICMR), for Senior Research Fellowship award to Prit Benny Malgulwar (No.3/2/3/284/2014/NCD-III).

## Supplementary Table

**Table-1:** List of primers used in the study

**Table-2:** Clinical and genetic features of ependymomas in the present study

## Supplementary Figure

**Supplementary Figure-1:** Expression analysis for BMI1 in 4 different ependymoma cohort (a) and on basis of different molecular subgroups as per GSE64415 (b). (c) Expression analysis for BMI1 on 1q status in *in-house* cohort

**Supplementary Figure-2:** (a) Expression analysis *FAL1* target genes in 4 gene expression microarray dataset of ependymomas. (b) CDKN1A/p21 levels in different molecular subgroups as per GSE64415. (c) Correlations between BMI1 and CDKN1A/p21 levels. (d) CDKN1A/p21 levels on 1q status in *in-house* cohort

**Supplementary Figure-3:** Kaplan-Meier curve for BMI1 and CDKN1A/p21 levels in GSE27287.

