## Supplementary Table-1 for "Focal amplification of *FAL1*, an oncogenic enhancer lncRNA mapping to chromosome 1q is associated with dysregulated BMI1/p21 axis and an adverse event in intracranial ependymomas"

**Supplementary Table 1 - List of primers used in study**

**a) FAL1 copy number gain**

| **Primers details** | |
| --- | --- |
| ***FAL1*** | FP-5’CCATGCTGGCACATGATCTA-3’ |
|  | FP-5’-TTCCCTCTGTGAAACCTGCT-3’ |
| **GAPDH** | FP- 5 ′-TCAAGAAGGTGGTGAAGCAG-3′ |
|  | RP- 5 ′-TGTCGCTGTTGAAGTCAG AG-3 |

**b) Gene expression analysis for FAL1, BMI1 and CDKN1A/p21**

| **Primers details** | |
| --- | --- |
| ***FAL1*** | FP- 5′-GCA AGC GGA GAC TTG TCT TT-3′ |
|  | RP- 5′-TTG AAC TCC TGA CCT CGT GA-3′ |
| **BMI1** | FP-5’- TCATCCTTCTGCTGATGCTG-3’ |
|  | RP-5’- CCGATCCAATCTGTTCTGGT-3’ |
| **CDKN1A/p21** | FP- 5′-CCT CAT CCC GTG TTC TCC TTT-3′ |
|  | RP-5′-GTA CCA CCC AGC GGA CAA GT-3′ |
| **GAPDH** | FP- 5′-GCC GTC TAG AAA AAC CTG CC-3 |
|  | RP-5′-ACC ACC TGG TGC TCA GTG TA-3′ |

**c) ChIP-qPCR**

| **Primers details** | |
| --- | --- |
| **CDKN1A/p21** | FP-5′ TCAGCCTACAGCACCTGTCA-3′ |
|  | RP-5′ CAGCAAGGCAGACAGAACAG- 3′ |
| **ACTB** | FP-5'-TCCCCTCCTTTTGCGAAAA-3' |
|  | RP- 5'-CGGCCAACGCCAAAACT-3' |

**d) RNA localization qPCR**

| **Primers details** | |
| --- | --- |
| ***FAL1*** | FP- 5′-GCA AGC GGA GAC TTG TCT TT-3′ |
|  | RP- 5′-TTG AAC TCC TGA CCT CGT GA-3′ |
| **U6** | CGCAAGGATGACACGCA |
| **GAPDH** | FP- 5′-GCC GTC TAG AAA AAC CTG CC-3 |
|  | RP-5′-ACC ACC TGG TGC TCA GTG TA-3′ |

**Supplementary Table 2 - Clinical and genetic features of ependymomas in the present study**

| Serial no | Age | Site | Grade | MIB status | 1q gain | *RELA* status | FAL1 status | FAL1 expression | BMI1 expression | p21 expression | Molecular Group |
| --- | --- | --- | --- | --- | --- | --- | --- | --- | --- | --- | --- |
| 1 | 1 | 2 | 3 | 1 | NA | NA | 1 | 1 | 1 | 0 | PFA |
| 2 | 1 | 1 | 3 | 1 | NA | Yes | 1 | 0 | 1 | 0 | RELA+ |
| 3 | 1 | 3 | 2 | 0 | NA | NA | 1 | 0 | 0 | 0 | SP |
| 4 | 2 | 3 | 2 | 0 | NA | NA | 0 | 0 | 0 | 1 | SP |
| 5 | 2 | 3 | 3 | 0 | NA | NA | 0 | 0 | 1 | 0 | SP |
| 6 | 1 | 2 | 3 | 0 | Yes | NA | 0 | 1 | 1 | 0 | PFA |
| 7 | 2 | 2 | 3 | 0 | No | NA | 1 | 1 | 1 | 0 | PFB |
| 8 | 2 | 2 | 2 | 0 | NA | NA | 1 | 1 | 1 | 1 | PFB |
| 9 | 2 | 3 | 2 | 0 | NA | NA | 0 | 1 | 0 | 0 | SP |
| 10 | 2 | 3 | 2 | 0 | NA | NA | 0 | 1 | 0 | 1 | SP |
| 11 | 1 | 3 | 2 | 0 | NA | NA | 0 | 1 | 1 | 1 | SP |
| 12 | 1 | 2 | 3 | 1 | No | NA | 0 | 0 | 0 | 1 | PFA |
| 13 | 1 | 1 | 3 | 0 | No | Yes | 1 | 1 | 1 | 0 | RELA+ |
| 14 | 1 | 2 | 3 | 0 | No | NA | 1 | 1 | 1 | 0 | PFA |
| 15 | 2 | 3 | 2 | 0 | NA | NA | 0 | 0 | 0 | 0 | SP |
| 16 | 1 | 1 | 2 | 0 | No | No | 1 | 1 | 1 | 0 | RELA- |
| 17 | 1 | 2 | 3 | 1 | No | NA | 1 | 1 | 1 | 0 | PFA |
| 18 | 2 | 1 | 3 | 0 | No | Yes | 1 | 1 | 0 | 0 | RELA+ |
| 19 | 1 | 2 | 3 | 1 | NA | NA | 1 | 1 | 1 | 0 | PFA |
| 20 | 1 | 1 | 3 | 1 | NA | Yes | 0 | 0 | 0 | 1 | RELA+ |
| 21 | 1 | 1 | 2 | 0 | NA | Yes | 1 | 1 | 1 | 0 | RELA+ |
| 22 | 1 | 2 | 3 | 1 | NA | NA | 1 | 1 | 1 | 0 | PFA |
| 23 | 1 | 3 | 3 | 1 | NA | NA | 0 | 1 | 0 | 1 | SP |
| 24 | 1 | 2 | 2 | 0 | Yes | NA | 0 | 0 | 0 | 1 | PFA |
| 25 | 1 | 2 | 3 | 0 | Yes | NA | 1 | 1 | 1 | 1 | PFA |
| 26 | 1 | 1 | 2 | 1 | No | No | 1 | 1 | 1 | 0 | RELA- |
| 27 | 1 | 1 | 3 | 1 | Yes | Yes | 1 | 1 | 1 | 0 | RELA+ |
| 28 | 2 | 2 | 2 | 0 | Yes | NA | 0 | 0 | 1 | 0 | PFB |
| 29 | 1 | 1 | 3 | 1 | Yes | Yes | 1 | 1 | 1 | 0 | RELA+ |
| 30 | 1 | 1 | 3 | 1 | Yes | Yes | 1 | 1 | 1 | 0 | RELA+ |
| 31 | 1 | 2 | 3 | 0 | No | NA | 0 | 1 | 1 | 0 | PFA |
| 32 | 1 | 1 | 3 | 0 | No | Yes | 0 | 1 | 1 | 0 | RELA+ |
| 33 | 1 | 2 | 2 | 0 | No | NA | 1 | 0 | 1 | 0 | PFA |
| 34 | 2 | 2 | 3 | 0 | Yes | NA | 1 | 1 | 0 | 0 | PFB |
| 35 | 1 | 1 | 3 | 0 | Yes | No | 0 | 0 | 1 | 0 | RELA- |
| 36 | 1 | 1 | 2 | 0 | NA | Yes | 1 | 1 | 1 | 0 | RELA+ |
| 37 | 2 | 1 | 3 | 0 | NA | No | 1 | 1 | 0 | 1 | RELA- |
| 38 | 2 | 1 | 3 | 0 | NA | No | 1 | 1 | 1 | 0 | RELA- |
| 39 | 2 | 1 | 3 | 1 | NA | Yes | 1 | 0 | 1 | 0 | RELA+ |
| 40 | 2 | 1 | 3 | 1 | NA | Yes | 1 | 0 | 0 | 0 | RELA+ |
| 41 | 1 | 1 | 3 | 1 | NA | Yes | 1 | 1 | 0 | 0 | RELA+ |
| 42 | 1 | 1 | 3 | 1 | NA | Yes | 1 | 1 | 1 | 1 | RELA+ |
| 43 | 1 | 1 | 3 | 1 | NA | Yes | 1 | 0 | 1 | 1 | RELA+ |
| 44 | 1 | 1 | 3 | 1 | NA | Yes | 0 | 0 | 1 | 0 | RELA+ |
| 45 | 1 | 1 | 3 | 1 | NA | No | 1 | 1 | 1 | 0 | RELA- |
| 46 | 2 | 1 | 2 | 0 | NA | No | 0 | 1 | 0 | 0 | RELA- |
| 47 | 1 | 1 | 3 | 1 | NA | Yes | 1 | 1 | 1 | 0 | RELA+ |

Age: <18 years-0, >18 years-1

Site: Supratentorial-1, Infratentorial-2, Spinal-3

MIB1 status: >10 LI- 1, <10 LI-0

FAL1 status: gain- 1, normal status-0

Expression of FAL1, BMI1 and CDKN1A/p21: High-1, Low-0

NA- Not available
