## Supplementary figures and images for "Focal amplification of *FAL1*, an oncogenic enhancer lncRNA mapping to chromosome 1q is associated with dysregulated BMI1/p21 axis and an adverse event in intracranial ependymomas"

### Supplementary Figure-1

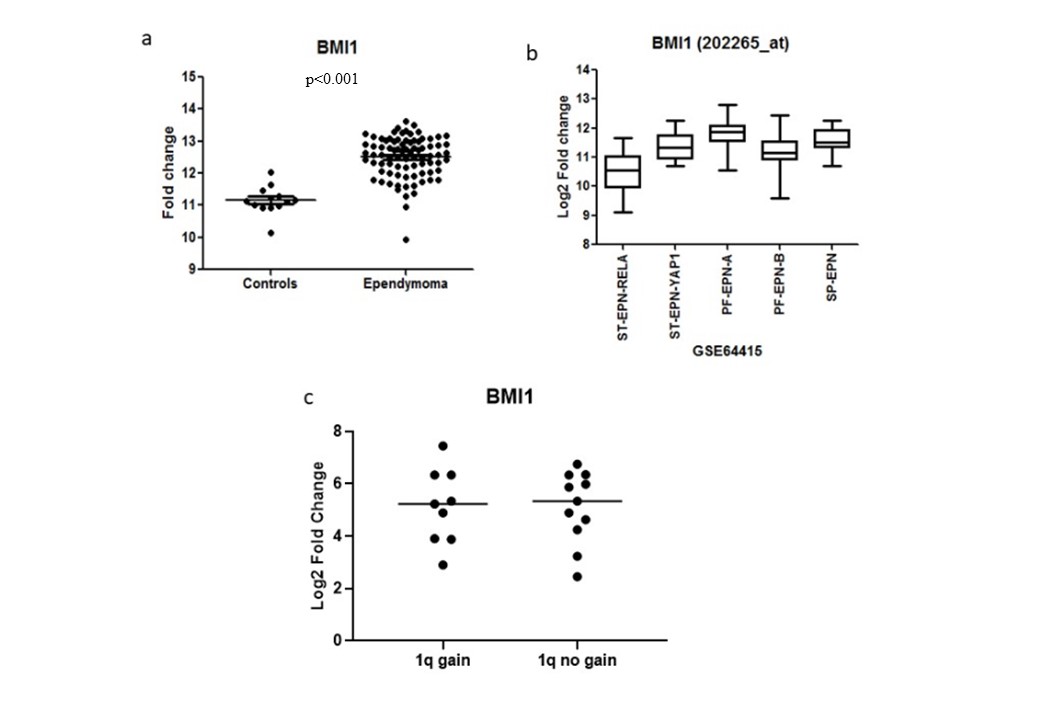

### Supplementary Figure-2

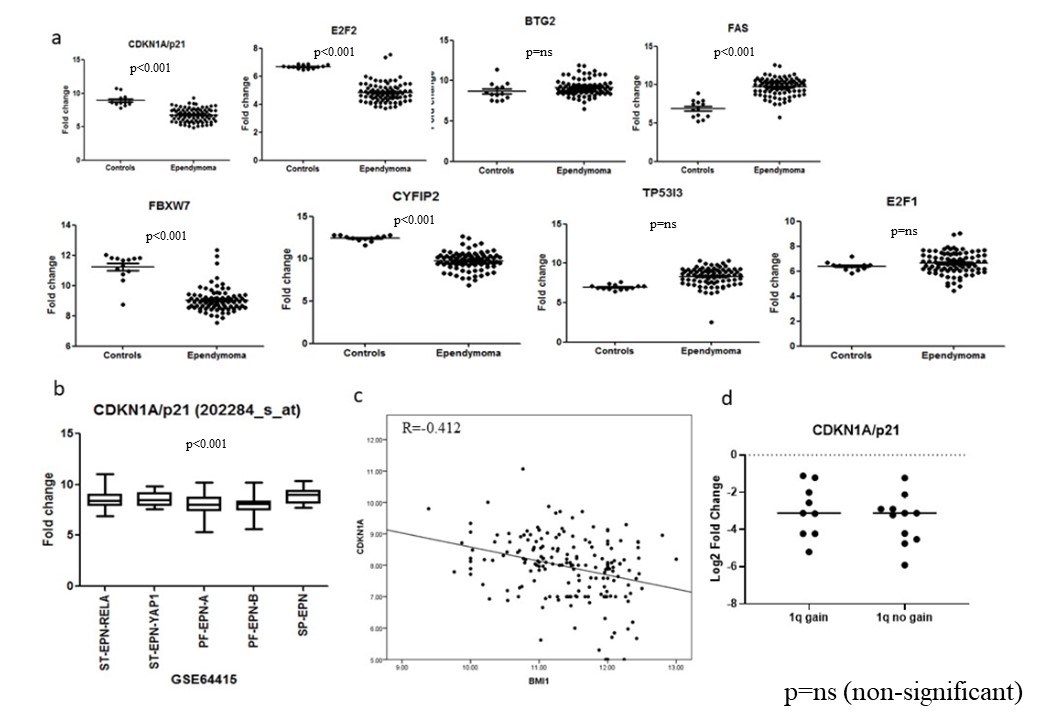

### Supplementary Figure-3

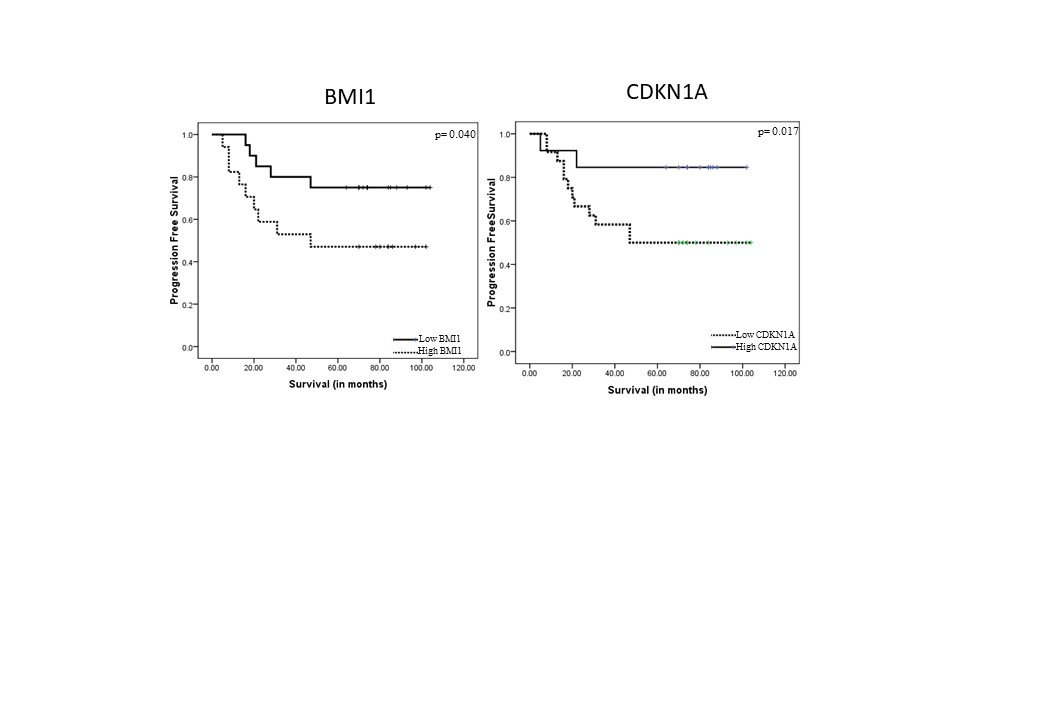
